# Proxalutamide Reduction of Mortality Rate in Hospitalized COVID-19 Patients Depends on Treatment Duration – an Exploratory Analysis of the Proxa-Rescue AndroCoV Trial

**DOI:** 10.1101/2021.06.28.21259661

**Authors:** Ricardo Ariel Zimerman, Daniel do Nascimento Fonseca, Michael do Nascimento Correia, Renan Nascimento Barros, Dirce Costa Onety, Karla Cristina Petruccelli Israel, Emilyn Oliveira Guerreiro, José Erique Miranda Medeiros, Raquel Neves Nicolau, Luiza Fernanda Mendonça Nicolau, Rafael Xavier Cunha, Maria Fernanda Rodrigues Barroco, Patrícia Souza da Silva, Raysa Wanzeller de Souza Paulain, Claudia Elizabeth Thompson, Andy Goren, Carlos Gustavo Wambier, Flávio Adsuara Cadegiani

## Abstract

**Introduction:** Proxalutamide, a second-generation non-steroidal antiandrogen (NSAA), primarily developed for castration-resistant prostate cancer, demonstrated reduction in 28-day mortality rate of 77.7% in hospitalized COVID-19 patients in a double-blind, placebo-controlled, two-arm randomized clinical trial (RCT), through intention-to-treat (ITT) analysis. We observed a high 28-day mortality rate of patients that did not complete the 14-day treatment with proxalutamide, compared to the placebo arm. These differences may raise hypotheses to explain the wide differences between ITT and on-treatment (OT) analysis in terms of efficacy. Despite the inherent limitations of OT analysis, we aimed to respond to unanswered questions regarding the drug efficacy when the 14-day treatment with proxalutamide was complete, and secondarily understand the causality relationship between treatment interruption and mortality rate.

**Methods:** This is a *post-hoc* exploratory analysis of a double-blinded, randomized, placebo-controlled, prospective, multicentric, two-arm RCT of 300mg-daily 14-day proxalutamide therapy for hospitalized COVID-19 patients not requiring mechanical ventilation. OT population excluded patients that did not complete the full 14-day course of therapy or died from COVID-19 complications within 24 hours of randomization. The primary outcome was the 28-day COVID-19 mortality rate. Secondary outcomes included median hospital length, 14-day and 28-day alive hospital discharge rate and 28-day all-cause mortality rate of those who discontinued intervention.

**Results:** In total, 580 patients completed the 14-day treatment or died during treatment, including 288 patients in the proxalutamide arm and 292 patients in the placebo arm, with similar baseline characteristics between groups. The 28-day COVID-19 mortality rate was 4.2% in the proxalutamide group and 49.0% in the placebo group. The mortality risk ratio (RR) was 0.08 (95% CI, 0.05-0.15), with a number needed to treat (NNT) of 2.2 to prevent death. The median hospital length stay after randomization was 5 days (interquartile range [IQR] = 3 to 7.2 days) in the proxalutamide group and 9 days (IQR = 6 to 15 days) in the placebo group (p <0.001). The 28-day all-cause mortality rate of patients that received proxalutamide but interrupted treatment before 14 days was 79.3%, while those that received placebo and interrupted before 14 days was 52.8% (p = 0.054 between groups).

**Conclusion:** The reduction in 28-day all-cause mortality rate with 14-day proxalutamide treatment for hospitalized COVID-19 patients was more significant while on treatment adhesion (92%), compared to the reduction when all patients enrolled in the proxalutamide arm were considered (77.7%). However, the magnitude of statistical significance of the reduction in all-cause mortality and the NNT were similar between the OT and ITT analysis. The apparent high mortality risk rate with early interruption of proxalutamide treatments suggests that strategies for treatment compliance should be reinforced for future RCTs with proxalutamide. (NCT04728802)

## Introduction

The identification of potential molecules that reduces severe acute respiratory syndrome coronavirus 2 (SARS-CoV-2) infectivity and pathogenicity and coronavirus disease 2019 (COVID-19) severity is critical while the COVID-19 pandemics is not fully controlled. While vaccines are considered as the most efficient public health measure to reduce COVID-19 mortality preventively, we still struggle to find effective drugs to reduce COVID-19 poorer outcomes among those already infected.^1^

The transmembrane protease serine 2 (TMPRSS2) is a key enzyme for viral cell entry, since it primes SARS-CoV-2 spike proteins to allow for a more efficient subsequent binding to membrane-attached angiotensin-converting enzyme 2 (ACE2) receptors.^1^ The only known endogenous modulators of TMPRSS2 expression are androgens. Antiandrogens are promising agents against COVID-19 due to their potential actions in the blockage of the entrance of the SARS-CoV-2 into the cells, through the inhibition of TMPRSS2 expression.^2,3^

Early observations in COVID-19 pandemic have identified hyperandrogenic phenotypes as independent risk factors for disease severity in both males ^4–6^ and females. ^7–9^ Antiandrogens have mechanistic plausibility to work against SARS-CoV-2,^10,11^ have demonstrated pre-clinical ^12–14^ and preliminary clinical efficacy against COVID-19, both when used continuously^15–17^ or initiated during the course of the disease.^18^

Proxalutamide is a second-generation non-steroidal antiandrogen (NSAA), primarily developed for castration-resistant prostate cancer, that demonstrated high potency as an androgen receptor (AR) antagonist. Proxalutamide also acts as a suppressor of AR gene expression and regulates the angiotensin converting enzyme-2 (ACE-2).^19^

We have previously demonstrating that proxalutamide was effective to prevent hospitalization in COVID-19 male patients and to reduce viral shedding and inflammatory response, in a double-blind, placebo-controlled randomized clinical trial (RCT).^20,21^

We have also demonstrated that proxalutamide was able to reduce mortality rate by 77.7% in moderate-to-severe hospitalized COVID-19 patients that were not on mechanical ventilation, and increased recovery speed by 128%, when compared to placebo, also in a double-blind, placebo-controlled RCT.

The efficacy of proxalutamide in hospitalized COVID-19 patients was demonstrated using intention-to-treat (ITT) analysis that did not require any type of adjustment. The unmodified ITT is the gold standard method of evaluation to demonstrate efficacy and is more conservative, since ITT analysis includes subjects that discontinued treatment.

Of the 645 patients enrolled in the trial, 65 discontinued or withdrew, including 29 of 317 subjects (9.1%) of the proxalutamide arm and 36 of 328 subjects (11.0%) of the placebo arm.^22^ The 28-day mortality rate was overwhelmingly high among those that interrupted proxalutamide (79.3% mortality rate - 23 out of 29 patients), while the mortality rate for those who interrupted placebo was 50.0% (18 out of 36 patients),^22^ similar to the observed in the overall placebo arm. Importantly, placebo non-compliant patients exhibited similar mortality rate when compared to both overall placebo arm of the trial and in-hospital mortality in the same places where the trial was conducted.^23^

The intriguing high mortality of patients that did not complete the proxalutamide treatment led us to hypothesize that drug efficacy when used appropriately for 14 days could be different from the efficacy observed in ITT analysis. To evaluate those subjects that completed treatment or that died during treatment only, an on-treatment (OT) analysis would be recommended, although it may not allow conclusions regarding the actual drug efficacy.

The objective of the present *post-hoc* exploratory analysis is to evaluate the efficacy of proxalutamide in COVID-19 hospitalized patients when 14-day treatment is completed or uninterrupted until death, in case death occurs before 14 days of treatment. In addition, we describe the consequences in terms of outcomes of shorter duration proxalutamide treatment.

## Methods

This is a *post-hoc* analysis of a double-blinded, randomized, placebo-controlled, prospective, two-arm RCT^22^ describing the results of the RCT restricted to OT analysis, *i*.*e*., excluding patients that discontinued treatment before 14 days of drug treatment.

A detailed description of the trial design, sample size calculation, settings, recruitment, patient selection criteria, randomization, blinding, procedures and statistical analysis are published in another manuscript and detailed elsewhere.^22^

The trial was conducted at eight centers in six cities of the state of Amazonas, Brazil. Patients were recruited between February 1 and March 17, 2021. The RCT was registered in clinicaltrials.gov (NCT04728802), and approved by Brazilian National Ethics Committee (Approval number #4.513.425; CAAE: 41909121.0.0000.5553; Comitê de Ética em Pesquisa (CEP) of the Comitê Nacional de Ética em Pesquisa (CONEP) of the Ministry of Health (MS)) (CEP/CONEP/MS).

### Selection criteria

Selection criteria included subjects hospitalized due to COVID-19, not in mechanical ventilation, with positive real-time reverse transcription polymerase chain reaction (rtPCR) (Roche, USA) for SARS-CoV-2 within seven days, and, upon randomization, without class III or IV congestive heart failure (New York Heart Association), immunosuppression, baseline alanine transferase (ALT) above five times ULN (> 250 U/L), creatinine above 2.5 mg/ml, and not currently using antiandrogen drugs, and, in case of women, not pregnant or planning to become pregnant within 90 days and nor breastfeeding.

### Procedures

Patients were randomized to receive either proxalutamide 300 mg/day plus usual care or a placebo plus usual care, for 14 days, in a 1:1 ratio.

The COVID-19 8-point ordinal scale was used to determine disease severity. The scale was employed during screening (day 0), on a daily basis on days 1-14, day 21, and day 28. The clinical score was defined as: 8. Death; 7. Hospitalized, on invasive mechanical ventilation; 6. Hospitalized, on non-invasive ventilation or high flow oxygen devices; 5. Hospitalized, requiring supplemental oxygen; 4. Hospitalized, not requiring supplemental oxygen-requiring ongoing medical care (COVID-19 related or otherwise); 3. Hospitalized, not requiring supplemental oxygen - no longer requires ongoing medical care; 2. Not hospitalized, limitation on activities; and 1. Not hospitalized, no limitations on activities.^24^

Patients discharged before the end of treatment continued the treatment at home until 14 days and were actively monitored for treatment compliance by daily phone calls. Participants discharged from the hospital were evaluated until day 28. Patients were instructed to contact or visit the same institution in case of relapse or new symptoms. Hospital readmissions were actively surveilled in all sites.

Baseline characteristics, previous medical history, and concomitant medications were recorded for each patient. Proxalutamide 300 mg/day (3 tablets of 100mg a day) or placebo (3 tablets a day) plus usual care was given for 14 days, even when COVID-19 remission occurred before this period. Usual care included medications such as enoxaparin, colchicine, methylprednisolone, dexamethasone, or antibiotic therapy if necessary.

### Outcomes

The primary outcome measure was the COVID-19 mortality rate at 28 days under OT analysis. The secondary endpoints included hospitalization length duration (days), the recovery rate, which was defined as achieving alive hospital discharge (scores 1 and 2) at day 14 and 28 under OT analysis. We also compared 28-day mortality rate between non-treatment completers of the proxalutamide *versus* placebo arm, as well as the differences between the present OT analysis and the ITT analysis, published elsewhere.^22^

### Statistical Analysis

Risk ratios (RR) and confidence interval (CI) were calculated for recovery and mortality rates, Cox proportional hazards model was used to calculate hazard ratio (HR) and CI for all-cause mortality at day 28, the Wilcoxon rank sum test was used for ordinal scale scores at 14 and 28 days, Kaplan-Meier’s survivor function was used to evaluate and illustrate mortality and recovery rates over the 28 days after randomization. Statistical significance was defined as P<0.05. Statistical analysis was performed in Stata/SE version 16.1 for Mac (StataCorp LLC, College Station, TX, USA).

## Results

### Subjects

In total, 580 patients completed the 14-day treatment or died during treatment. Of these, 288 patients were treated with proxalutamide, including 163 males (56.7%) and 125 females (43.3%), and 292 patients received placebo, including 166 males (56.8%) and 126 females (43.2%). This population was considered for the OT analysis. A chart describing the patient flow of the OT population study is shown in **Figure 1**.

**Figure 1.**
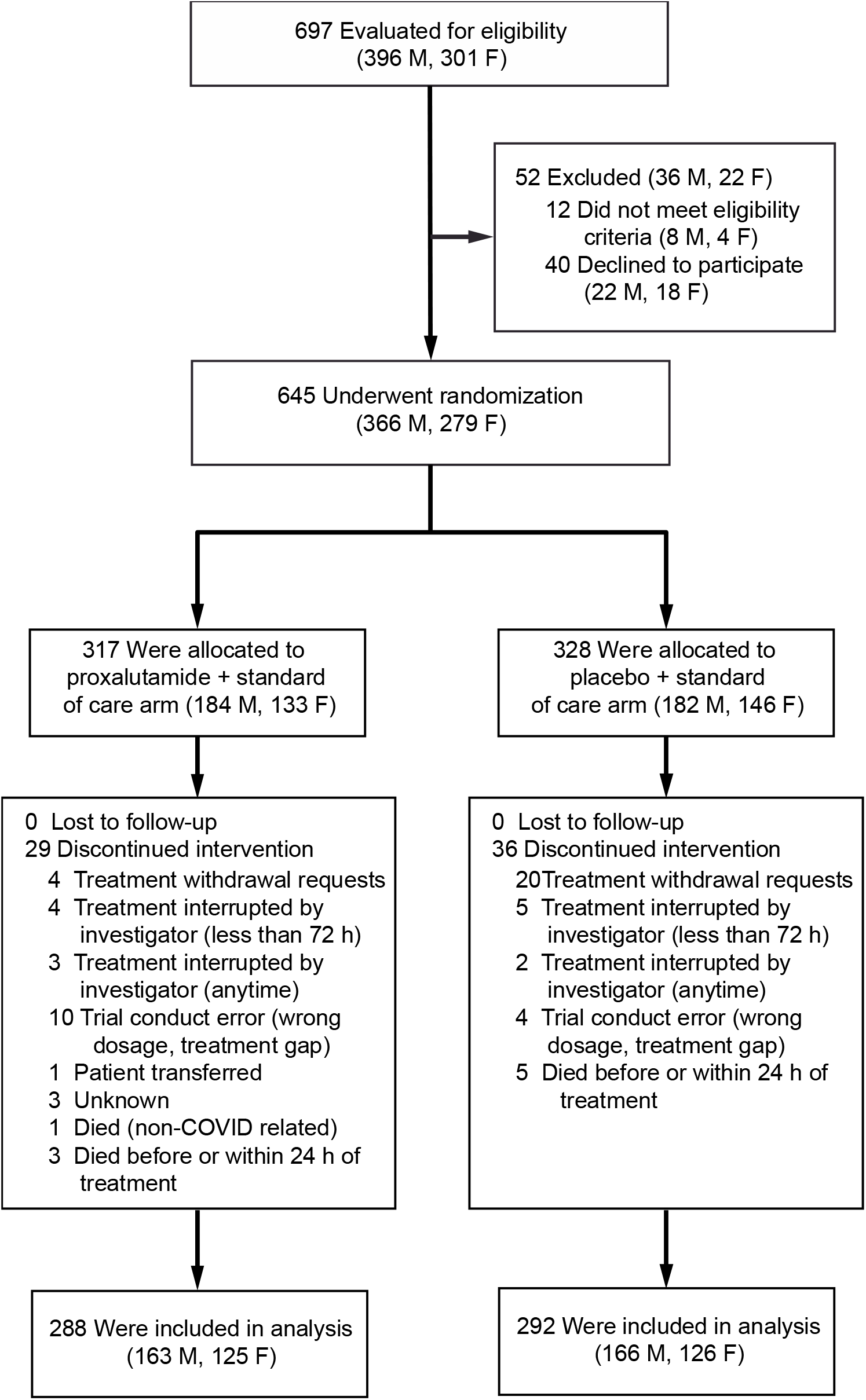
Enrollment, randomization, and analysis of the on-treatment (OT) population.

The baseline characteristics of the study population OT are described in **Table 1**. The median age was 50.0 years (inter-quartile range, 39.0 to 61.0) Hypertension, type 2 diabetes mellitus (T2DM) and chronic obstructive pulmonary disorder (COPD) were present in 25.1%, 11.7%, and 2.1% of the patients, and the body mass index (BMI) was above 30 in 8.0% of patients. No comorbidities, one comorbidity, and two or more comorbidities were present in 69.2%, 17.1%, and 13.1%, of the patients, respectively. Median time from hospitalization to randomization was 2.0 days (inter-quartile range, 1.0 to 4.0). The ordinal scale score at baseline were 6 (hospitalized on high-flow oxygen or non-invasive ventilation), 5 (hospitalized with oxygen use), and 4 (hospitalized with no oxygen use, but requiring medical care) in 66.4%, 30.5%, and 2.7% of the patients, respectively. Median age, prevalence of comorbidities, number of comorbidities, median time from hospitalization to randomization, and distribution of ordinal scale score were similar between proxalutamide and placebo groups, and were also similar between per protocol and intent-to-treat cohorts.

**Table 1.**
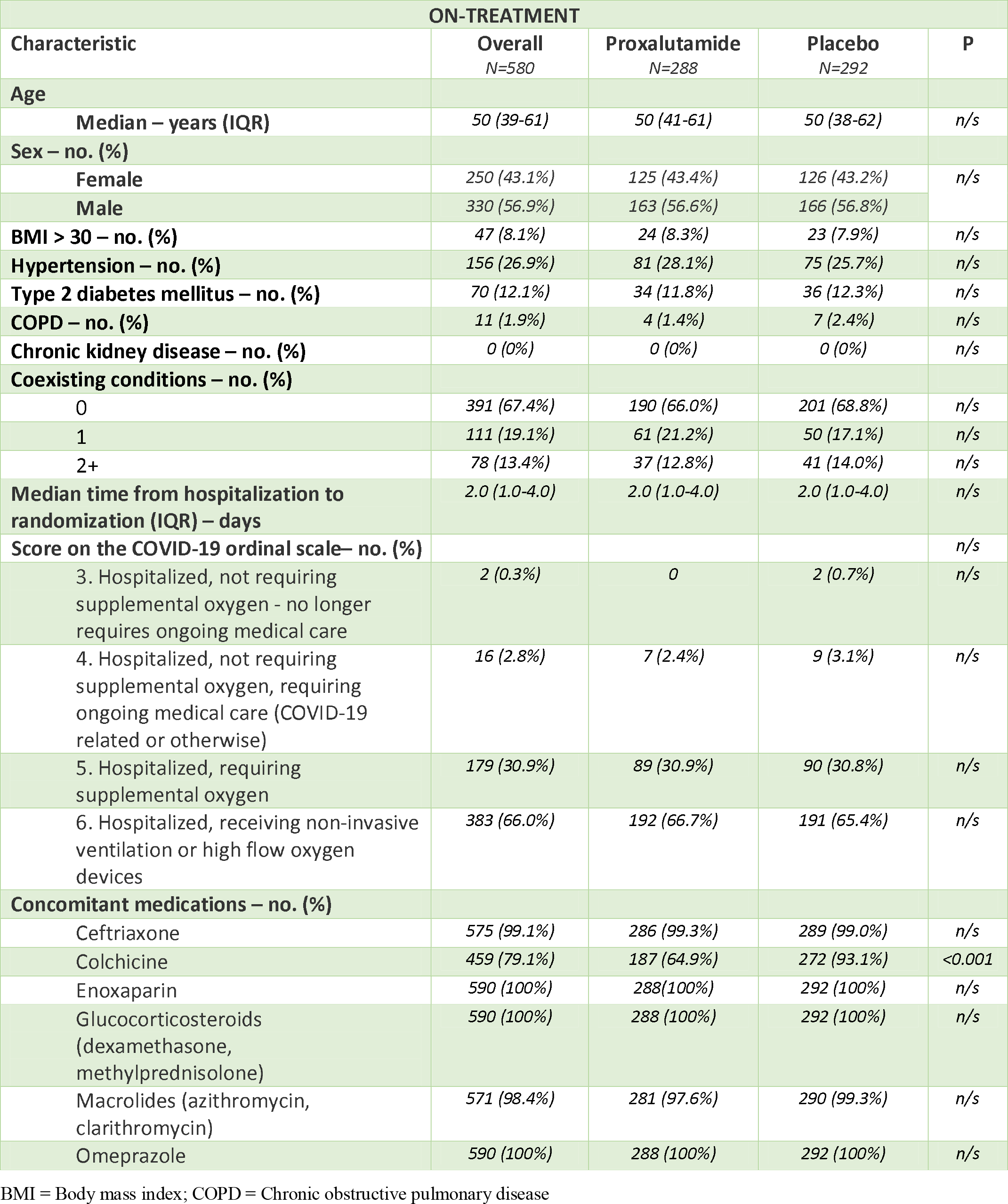
On-treatment baseline clinical characteristics, comorbidities, and concomitant medications.

Enoxaparin, omeprazole were administered to 100% of patients. Ceftriaxone, macrolides (azithromycin, clarithromycin) and colchicine were given to 99.3%, 97.6%, and 64.8% of patients receiving proxalutamide, and 99.0%, 99.3%, and 92.6% of patients receiving placebo, respectively. All concomitant medications were used at similar proportions between the groups, except for colchicine (p <0.001).

### Efficacy Outcomes

Table 2 describes the primary and secondary outcomes OT, and **Figure 2** illustrates the Kaplan-Meier curves of the outcomes from randomization until day 28.

The 28-day COVID-19 mortality rates were 4.2% for the proxalutamide arm and 49% for the placebo arm. The 28-day COVID-19 mortality ratio was 0.08 (95% CI, 0.05-0.15) at day 28.

**Table 2.**
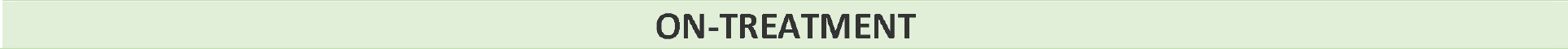
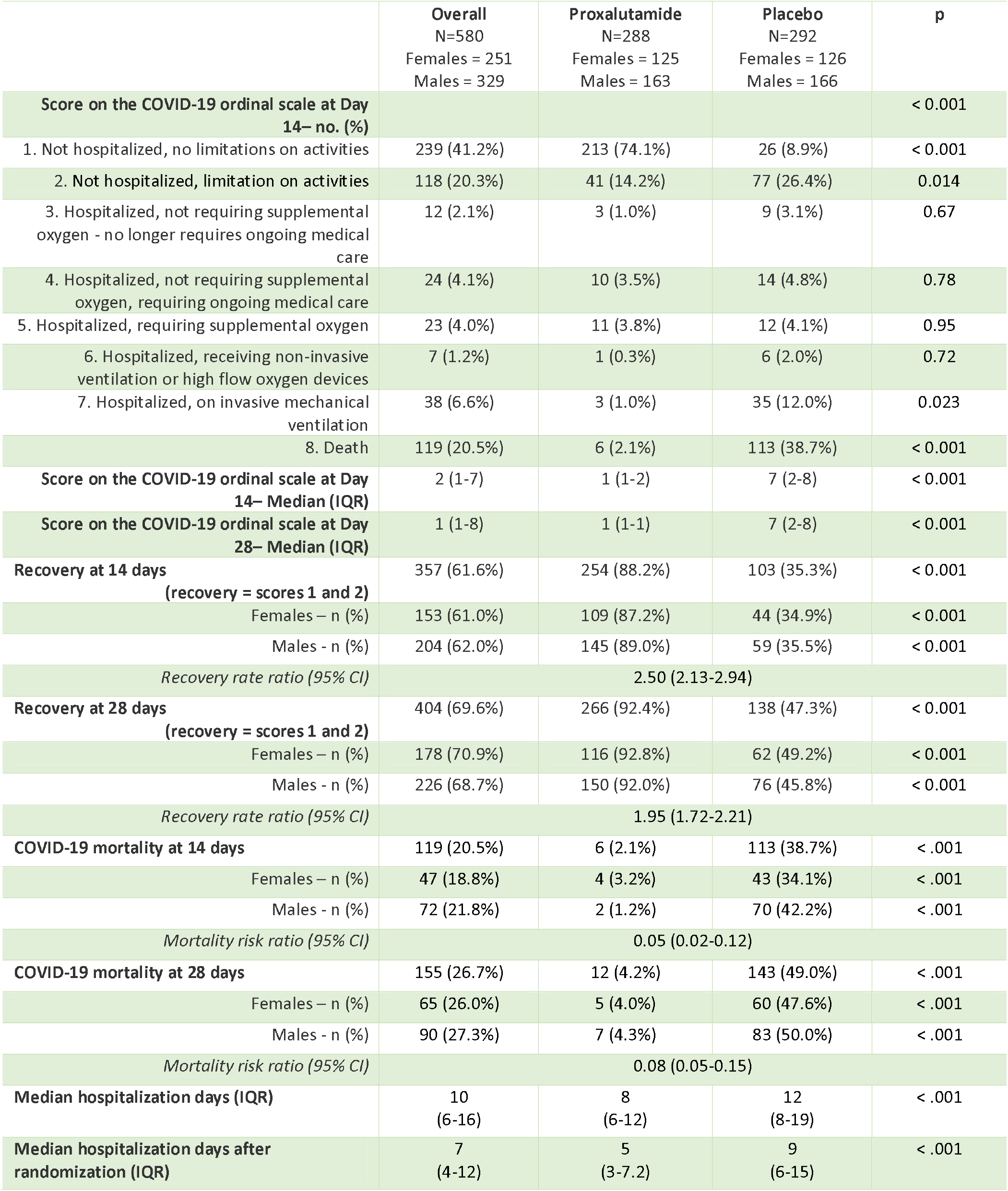
Primary and secondary outcomes, on-treatment analysis. Coronavirus disease 2019 8-point ordinal scale scores distribution at 14 days. Mortality (score 8), and recovery rates (scores 1 or 2) over 14 and 28 days after randomization (IQR = interquartile range, CI = confidence interval).

**Figure 2.**
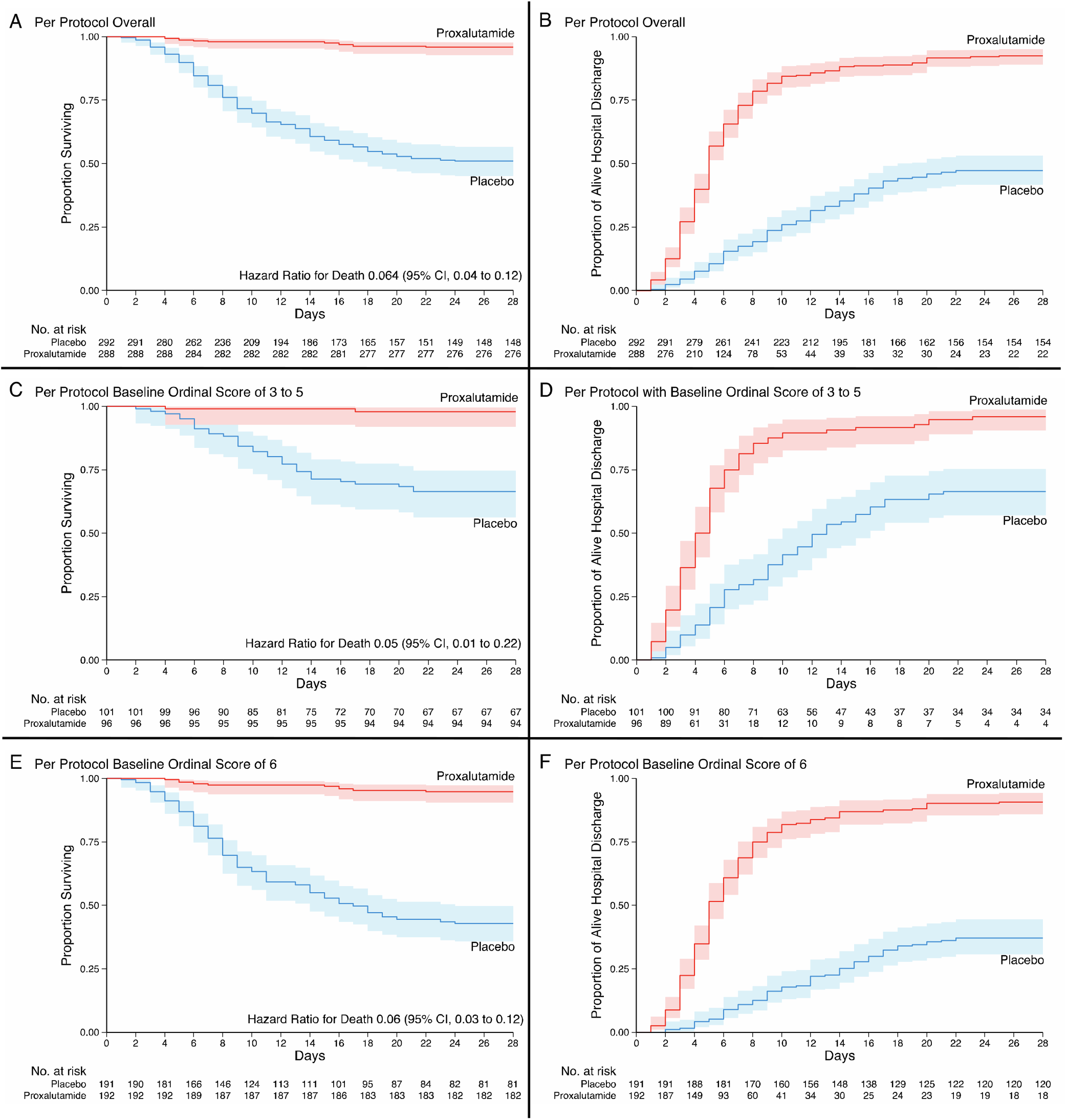
Kaplan–Meier estimates in the on-treatment analysis from randomization to Day 28. Surviving (Panel A) and Alive Hospital Discharge (Panel B) for the overall population. Surviving (Panel C) and Alive Hospital Discharge (Panel D) for patients with baseline ordinal scale scores 3 to 5. Surviving (Panel E) and Alive Hospital Discharge (Panel F) for patients with baseline ordinal scale score 6.

The median hospital length stay was eight days (IQR, 6 to 12) in the proxalutamide group and 12 days (IQR, 8 to 19) in the placebo group (p < 0.001). When considering the number of hospitalization days since beginning of treatment, median duration was five (IQR, 3 to 7.2) and nine days (IQR, 6 to 15) in the proxalutamide and placebo groups, respectively.

After 14 days, distribution of disease burden was significantly different between proxalutamide and placebo (p < 0.001). Among 288 patients from the proxalutamide group, six (2.1%) died until day 14, and 113 (38.7%) of the 292 patients from the placebo group died by day 14 (p < 0.001). At day 28, 12 (4.2%) and 143 (49.0%) patients died from the proxalutamide and placebo groups, respectively (p < 0.001). The mortality risk ratio was 0.05 (95% CI, 0.02-0.12) at day 14.

Recovery rates were 88.2% and 92.4% in the proxalutamide group at days 14 and 28, and 35.3% and 47.3% in the placebo group, respectively. The recovery rate ratio was 2.50 (95%CI, 2.13-2.94) at day 14 and 1.95 (95% CI, 1.72-2.21) at day 28, favorable for proxalutamide *versus* placebo.

The 28-day all-cause mortality rate of patients receiving proxalutamide that interrupted the 14-day proxalutamide treatment at least 24 hours before death (79.3%; 23 deaths of 29 patients) was marginally significantly higher than the 28-day mortality rate of non-compliant patients that were receiving placebo (52.8%; 19 deaths of 36 patients) (p = 0.058).

### Safety Outcomes

Adverse events (AEs) observed in the population OT are detailed in Table 3. Number of subjects that experimented at least one adverse effect was higher in the placebo group (64.4%) than in the proxalutamide group (26.0%) (p < 0.001). Disease progression was more commonly observed in the placebo arm (51.7%) than in the proxalutamide arm (4.5%) (p < 0.001). Shock needing use of vasopressors was more common in the placebo group (42.8%) than in the proxalutamide group (1.4%) (p < 0.001). Liver or kidney injury was more commonly detected in the placebo group (12.0%) than in the proxalutamide group (1.7%) (p = 0.032). Among grades 2 and 1 AEs, diarrhea was the only AEs more commonly reported in the proxalutamide arm (16.3%) than in the placebo arm (3.1%) (p = 0.006).

**Table 3.**
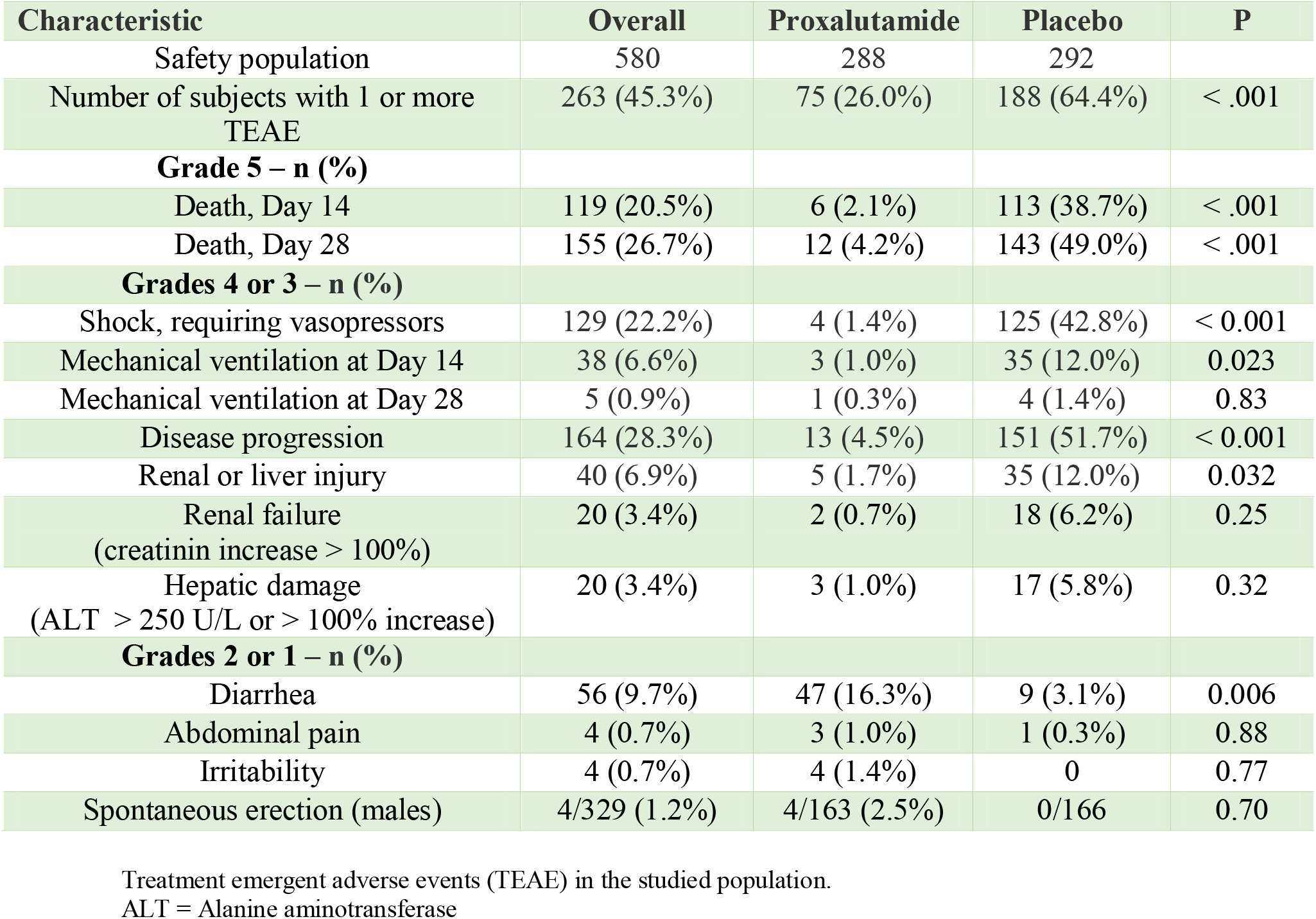
On-treatment safety outcomes.

| Characteristic | Overall | Proxalutamide | Placebo | P |
| --- | --- | --- | --- | --- |
| Safety population | 580 | 288 | 292 |  |
| Number of subjects with 1 or more TEAE | 263 (45.3%) | 75 (26.0%) | 188 (64.4%) | < .001 |
| **Grade 5 – n (%)** |  |  |  |  |
| Death, Day 14 | 119 (20.5%) | 6 (2.1%) | 113 (38.7%) | < .001 |
| Death, Day 28 | 155 (26.7%) | 12 (4.2%) | 143 (49.0%) | < .001 |
| **Grades 4 or 3 – n (%)** |  |  |  |  |
| Shock, requiring vasopressors | 129 (22.2%) | 4 (1.4%) | 125 (42.8%) | < 0.001 |
| Mechanical ventilation at Day 14 | 38 (6.6%) | 3 (1.0%) | 35 (12.0%) | 0.023 |
| Mechanical ventilation at Day 28 | 5 (0.9%) | 1 (0.3%) | 4 (1.4%) | 0.83 |
| Disease progression | 164 (28.3%) | 13 (4.5%) | 151 (51.7%) | < 0.001 |
| Renal or liver injury | 40 (6.9%) | 5 (1.7%) | 35 (12.0%) | 0.032 |
| Renal failure<br>(creatinin increase > 100%) | 20 (3.4%) | 2 (0.7%) | 18 (6.2%) | 0.25 |
| Hepatic damage<br>(ALT > 250 U/L or > 100% increase) | 20 (3.4%) | 3 (1.0%) | 17 (5.8%) | 0.32 |
| **Grades 2 or 1 – n (%)** |  |  |  |  |
| Diarrhea | 56 (9.7%) | 47 (16.3%) | 9 (3.1%) | 0.006 |
| Abdominal pain | 4 (0.7%) | 3 (1.0%) | 1 (0.3%) | 0.88 |
| Irritability | 4 (0.7%) | 4 (1.4%) | 0 | 0.77 |
| Spontaneous erection (males) | 4/329 (1.2%) | 4/163 (2.5%) | 0/166 | 0.70 |

Treatment emergent adverse events (TEAE) in the studied population. ALT = Alanine aminotransferase

## Discussion

Reduction of mortality with the use of proxalutamide 300mg daily for 14 days in hospitalized COVID-19 patients was not-negligently higher in the OT analysis (95% and 92% reduction at days 14 and 28, respectively) than in the ITT (79% and 78% reduction at days 14 and 28, respectively).^22^ The differences between ITT and OT occurred because the 28-day mortality rate of treatment non-completers, *i*.*e*., patients that did not complete the 14-day proxalutamide treatment or that died at least 24 hours before interruption, was extremely high (79.3%), and substantially higher than non-compliant patients that were receiving placebo (52.8%) and fully-compliant that received proxalutamide (4.2%). We hypothesized that the sudden and early interruption of proxalutamide may have led to an overcompensating relapse of COVID-19 and its complications. The mortality rate in non-treatment compliance subjects reinforces the antiviral and indirect protective mechanisms of action of proxalutamide, as a sort of proof-of-concept, since its early removal was mostly fatal for patients. This also raises concern regarding the importance to comply to the period of use of 14 days.

Despite the important differences between the OT and ITT analysis, the NNT of proxalutamide for hospitalized COVID-19 patients to prevent death was numerically similar between OT and ITT analysis, of 2.2 and 2.6, respectively. The interpretation of the results of proxalutamide as being highly effective to prevent deaths in moderate-to-severe COVID-19 patients did not change between ITT and OT analyses.

This study has limitations inherent to *post-hoc* and retrospective analyses, although all the major barriers that could lead to confounding biases were addressed.^25^ Unbalanced actives and placebo distribution across sites and minor protocol modifications were the major weaknesses of the present RCT, while the lack of any modification of the ITT analysis prevented additional biases.^21^

The major confounding factor in the present analysis is the causality relationship between interrupting of treatment with proxalutamide and apparent higher mortality rate. Initially, differences in the mortality rate observed between the OT and ITT analysis could be due to the fact that patients tend to discontinue treatment in case of lack of response. Since the mortality rate was substantially lower in the proxalutamide arm compared to the placebo arm, and the number of non-completers was similar between these groups, similar increases in mortality rate would represent a higher increase in terms of percentage in the group with lower mortality rate, *i*.*e*., in the active arm.

However, treatment non-compliance in the placebo arm was not associated to poorer response compared to completers of the placebo arm. Conversely, non-completers of the proxalutamide arm had higher mortality rate compared to completers of the same arm and to the placebo arm. This reinforces the hypothesis of potential harm caused by abrupt and early interruption of proxalutamide.

Indeed, in the male outpatient trial with proxalutamide,^20^ the initial protocol was a 3-day treatment. However, after a few cases of disease relapse after the discontinuation, we increased the treatment duration to 7 days. In addition, in this trial, all 3 hospitalizations occur only after the end of the treatment with proxalutamide.

We reinforce the fact that the high mortality rate observed in non-compliance patients raised questions regarding the early interruption of proxalutamide, not only as an issue of no longer providing protection against SARS-CoV-2, but also the early discontinuation as being harmful and dangerous when duration of treatment is not respected. Since it is largely known that duration of a drug therapy is highly critical for its effects and outcomes, the comparisons between OT and ITT analysis and between early interruption of proxalutamide *versus* early interruption of placebo was necessary to highlight the importance of treatment duration with proxalutamide for moderate-to-severe COVID-19 explicit.

In addition to the message that the actual reduction of mortality rate with proxalutamide when treatment was fully complied is 92%, the present work also conveys the message that further trials with proxalutamide must include in their consent form that a potential increased risk of mortality if treatment is interrupted before the proposed period of 14 days. This warning is of great ethics importance and is made necessary for any future study on the drug. Ongoing trials should consider amending their consent forms, warn enrolled patients of these potential increased risk of mortality, and ethics committees should be informed of this new finding for our RCT.

## Data Availability

Data is available in case request is approved by the research team.

